# A mathematical model for the spatiotemporal epidemic spreading of COVID19

**DOI:** 10.1101/2020.03.21.20040022

**Authors:** Alex Arenas, Wesley Cota, Jesús Gómez-Gardeñes, Sergio Gómez, Clara Granell, Joan T. Matamalas, David Soriano, Benjamin Steinegger

## Abstract

An outbreak of a novel coronavirus, named SARS-CoV-2, that provokes the COVID-19 disease, was first reported in Hubei, mainland China on 31 December 2019. As of 20 March 2020, cases have been reported in 166 countries/regions, including cases of human-to-human transmission around the world. The proportions of this epidemics is probably one of the largest challenges faced by our interconnected modern societies. According to the current epidemiological reports, the large basic reproduction number, *R*_0_ ∼ 2.3, number of secondary cases produced by an infected individual in a population of susceptible individuals, as well as an asymptomatic period (up to 14 days) in which infectious individuals are undetectable without further analysis, pave the way for a major crisis of the national health capacity systems. Recent scientific reports have pointed out that the detected cases of COVID19 at young ages is strikingly short and that lethality is concentrated at large ages. Here we adapt a Microscopic Markov Chain Approach (MMCA) metapopulation mobility model to capture the spread of COVID-19. We propose a model that stratifies the population by ages, and account for the different incidences of the disease at each strata. The model is used to predict the incidence of the epidemics in a spatial population through time, permitting investigation of control measures. The model is applied to the current epidemic in Spain, using the estimates of the epidemiological parameters and the mobility and demographic census data of the national institute of statistics (INE). The results indicate that the peak of incidence will happen in the first half of April 2020 in absence of mobility restrictions. These results can be refined with improved estimates of epidemiological parameters, and can be adapted to precise mobility restrictions at the level of municipalities. The current estimates largely compromises the Spanish health capacity system, in particular that for intensive care units, from the end of March. However, the model allows for the scrutiny of containment measures that can be used for health authorities to forecast with accuracy their impact in prevalence of COVID–19. Here we show by testing different epidemic containment scenarios that we urge to enforce total lockdown to avoid a massive collapse of the Spanish national health system.

## I. INTRODUCTION

As of 20 March 2020 the outbreak of the novel coronavirus, SARS-CoV-2, has infected more than 270.000 persons worldwide with COVID-19, killing more than 11.300. Epidemiological analysis of the outbreak have been used to estimate epidemiologically relevant parameters [1–10], and available mathematical models have been used to track and anticipate the spread of the epidemics [11–17]. Nevertheless, the particularities of the current epidemics calls for a rethinking of conventional models towards tailored ones. Here, we propose mathematical model particularly designed to capture the main ingredients characterising the propagation of SARS-CoV-2 and the clinical characteristics reported for the cases of COVID-19. To this aim, we rely on previous metapopulation models by the authors [18–21] including the spatial demographical distribution and recurrent mobility patterns, and develop a more refined epidemic model that incorporates the stratification of population by age in order to consider the different epidemiological and clinical features associated to each group age that have been reported so far. The mathematical formulation of these models rely on the Microscopic Markov Chain Approach formulation for epidemic spreading in complex networks [22–27].

The epidemic model we propose takes into account several specific characteristics of the dynamics of COVID-19, such as the important effect of asymptomatic (or with mild symptoms) infectious individuals, which may explain the large incidence of the epidemics. We also consider the fraction of individuals which require hospitalization to ICU, since their saturation constitutes one of the major political and health problems of COVID-19 outbreak. The result is a model with seven epidemiological compartments for each of the patches composing the metapopulation. Additionally, we split the former epidemiological partition into three age groups: young, adults, and elderly people. This partition allows us to capture in a stylised way the main epidemiological, clinical and behavioural differences between the groups. On one hand, SARS-CoV-2 importation and exportation events between patches are mostly due to the mobility of active population. On the other hand, the medical evolution of COVID-19 displays strong differences across age groups [13, 28, 29]. In this regard, infections in the young group lead to mild symptoms that, without test, are often confused with those of a common cold, whereas for old individuals the infection evolves towards more severe symptoms and usually requires hospitalization.

The model incorporates the possibility of designing and evaluating the impact of contention policies to stop the propagation of SARS-CoV-2. In particular, we focus on those policies relying on global or targeted quarantine measures. They allow the selection of the optimum degree of mobility to avoid the health system crisis. Taking advantage of this possibility, we explore several epidemic scenarios characterized by different contention measures promoted on March 20, and evaluate their impact on the decrease of the epidemic prevalence and the saturation of the Spanish health system.

In a nutshell, the proposed model takes into account in an stylized way three main ingredients taking place in SARS-CoV-2 transmission: (i) the silent transmission of the pathogen through the young portion of the population, (ii) the large potential for the spatial dissemination of the pathogen provided by the mobility of mature individuals, and (iii) the severe symptoms caused of COVID-19 in elderly that yields to a dramatic increase of medical and hospital demands. Thus, the model can be viewed as three coevolving spreading processes with different spatio-temporal scales.

## II. EPIDEMIC SPREADING MODEL

We propose a tailored model for the epidemic spread of COVID-19. We use a previous framework for the study of epidemics in structured metapopulations, with heterogeneous agents, subjected to recurrent mobility patterns [18–20, 31].To understand the geographical diffusion of the disease, as a result of human-human interactions in small geographical patches, one has to combine the contagion process with the long-range disease propagation due to human mobility across different spatial scales. For the case of epidemic modeling, the metapopulation scenario is as follows. A population is distributed in a set of patches, being the size (number of individuals) of each patch in principle different. The individuals within each patch are well-mixed, *i*.*e*., pathogens can be transmitted from an infected host to any of the healthy agents placed in the same patch with the same probability. The second aspect of our metapopulation model concerns the mobility of agents. Each host is allowed to change its current location and occupy another patch, thus fostering the spread of pathogens at the system level. Mobility of agents between different patches is usually represented in terms of a network where nodes are locations while a link between two patches represents the possibility of moving between them.

We introduce a set of modifications to the standard metapopulation model to account for the different states relevant for the description of COVID-19, and also to substitute the well-mixing with a more realistic set of contacts. Another key point is the introduction of a differentiation of the course of the epidemics that depends on the demographic ages of the population. This differentiation is very relevant in light of the observation of a scarcely set of infected individuals at ages (< 25), and also because of the severe situations reported for people at older ages (> 65). Our model is composed of the following epidemiological compartments: susceptible (S), exposed (E), asymptomatic infectious (A), infected (I), hospitalized to ICU (H), dead (D), and recovered (R). Additionally, we divide the individuals in *N*_*G*_ age strata, and suppose the geographical area is divided in *N* regions or patches. Although we present the model in general form, its application to COVID-19 only makes use of the three age groups mentioned above (*N*_*G*_ = 3): young people (Y), with age up to 25; adults (M), with age between 26 and 65; and elderly people (O), with age larger than 65. See Figure 1 for an sketch of the compartmental epidemic model proposed.

**FIG 1.**
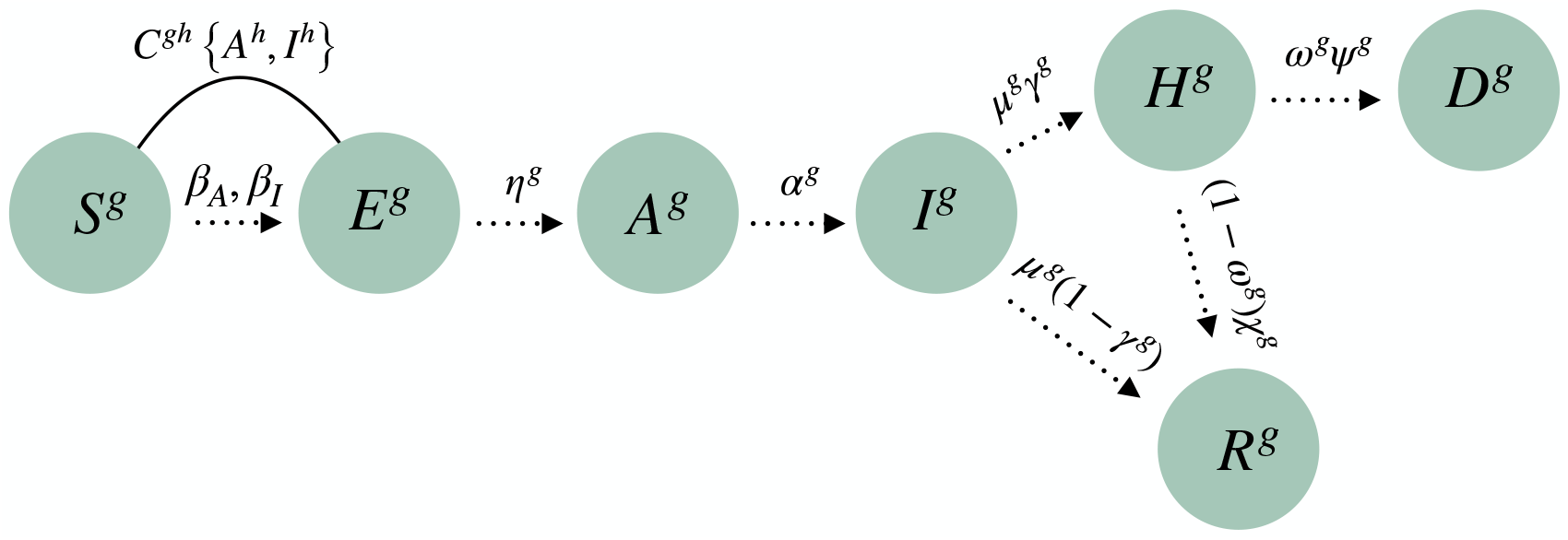
Compartemental epidemic model proposed in this study. The acronyms are susceptible (*S*^*g*^), exposed (*E*^*g*^), asymptomatic infectious (*A*^*g*^), infected (*I*^*g*^), hospitalized to ICU (*H*^*g*^), dead (*D*^*g*^), and recovered (*R*^*g*^), where g denotes for all cases the age stratum.

We characterize the evolution of the fraction of agents in state *m* ∈ {*S, E, A, I, H, D, R*} and for each age stratum *g* ∈ {1, …, *N*_*G*_}, associated with each patch *i* ∈ {1, …, *N*}, denoted in the following as 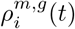 The temporal evolution of these quantities is given by:

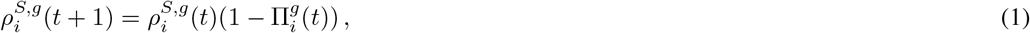

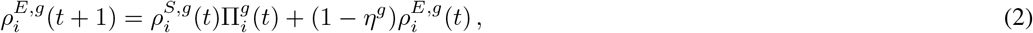

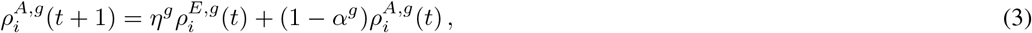

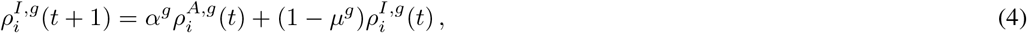

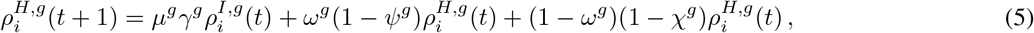

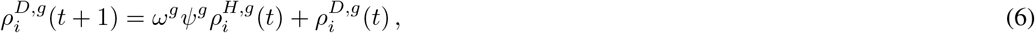

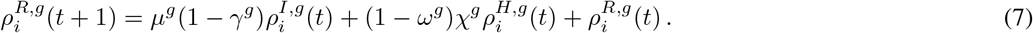

These equations correspond to a discrete-time dynamics, in which each time-step represents a day. They are built upon previous work on Microscopic Markov-Chain Approach (MMCA) modelization of epidemic spreading dynamics [22], but which has also been applied to other types of processes, *e*.*g*., information spreading and traffic congestion [24, 25, 32].

The rationale of the model is the following. Susceptible individuals get infected by contacts with asymptomatic and infected agents, with a probability 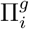, becoming exposed. Exposed individuals turn into asymptomatic at a certain rate *η*^*g*^, which in turn become infected at a rate *α*^*g*^. Once infected, two paths emerge, which are reached at an escape rate *µ*^*g*^. The first option is requiring hospitalization in an ICU, with a certain probability *γ*^*g*^; otherwise, the individuals become recovered. While being at ICU, individuals have a death probability *ω*^*g*^, which is reached at a rate *Ψ*^*g*^. Finally, ICUs discharge at a rate *χ*^*g*^, leading to the recovered compartment. See Table I for a summary of the parameters of the model, and their values to simulate the spreading of COVID-19 in Spain, which will be discussed in Subsec. IV A.

**TABLE I.** Parameters of the model and their estimations for COVID-19. See section IV.a for a detailed explanation.

| Symbol | Description | COVID-19 estimations for age groups $g \in \{Y, M, O\}$ in Spain |
| --- | --- | --- |
| $\beta_A$ | Infectivity of asymptomatic | 0.06 |
| $\beta_I$ | Infectivity of infected | 0.06 |
| $\langle k^g \rangle$ | Average number of contacts | (11.8, 13.3, 6.6) |
| $\eta^g$ | Latent rate | $\frac{1}{2.34}$ |
| $\alpha^g$ | Asymptomatic infectious rate | $\left( \frac{1}{5.06}, \frac{1}{2.86}, \frac{1}{2.86} \right)$ |
| $\mu^g$ | Escape rate | $\left( \frac{1}{1.0}, \frac{1}{3.2}, \frac{1}{3.2} \right)$ |
| $\gamma^g$ | Fraction of cases requiring ICU | (0.002, 0.05, 0.36) |
| $\omega^g$ | Fatality rate of ICU patients | 0.42 |
| $\psi^g$ | Death rate | $\frac{1}{7.0}$ |
| $\chi^g$ | ICU discharge rate | $\frac{1}{10.0}$ |
| $n_i^g$ | Regional population | Data provided by INE |
| $R_{ij}^g$ | Mobility matrix | Data provided by INE |
| $C^{gh}$ | Contacts-by-age matrix | $\begin{pmatrix} 0.5980 & 0.3849 & 0.0171 \\ 0.2440 & 0.7210 & 0.0350 \\ 0.1919 & 0.5705 & 0.2376 \end{pmatrix}$ |
| $\xi$ | Density factor | 0.01 |
| $p^g$ | Mobility factor | (0.0, 1.0, 0.0) |
| $\sigma$ | Average household size | 2.5 |
| $\kappa_0$ | Confinement factor | Adjustable for containment |

The value of 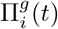 encodes the probability that a susceptible agent belonging to age group *g* and patch *i* contracts the disease. Under the model assumptions, this probability is given by:

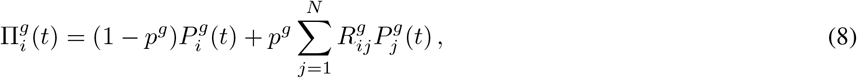

where *p*^*g*^ denotes the degree of mobility of individuals within age group *g*, and 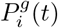 denotes the probability that those agents get infected by the pathogen inside patch *i*. This way, the first term in the r.h.s. of Eq. (8) denotes the probability of contracting the disease inside the residence patch, whereas the second term contains those contagions taking place in any of the neighboring areas. Furthermore, we assume that the number of contacts increases with the density of each area according to a monotonously increasing function *f*. Finally, we introduce an age-specific contact matrix, *C*, whose elements *C*^*gh*^ define the fraction of contacts that individuals of age group *g* perform with individuals belonging to age group *h*. With the above definitions, 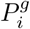 reads

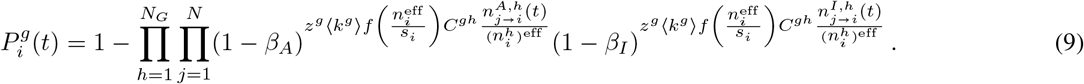

The exponents represent the number of contacts made by an agent of age group *g* in patch *i* with infectious individuals — compartments *A* and *I*, respectively — of age group *h* residing at patch *j*. Accordingly, the double product expresses the probability for an individual belonging to age group *g* not being infected while staying in patch *i*.

The term 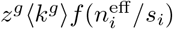 in Eq. (9) represents the overall number of contacts (infectious or non infectious), which increases with the density of patch *i* following function *f*, and also accounts for the normalization factor *z*^*g*^, which is calculated as:

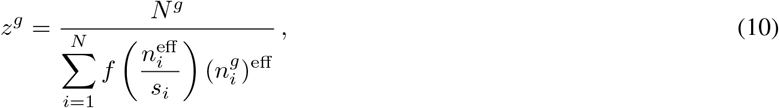

where the effective population at patch *i* is given by

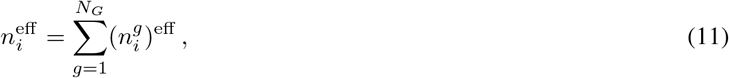

which is distributed in age groups of size

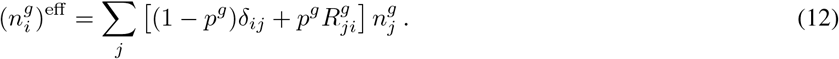

The function *f* (*x*) governing the influence of population density has been selected, following [33], as:

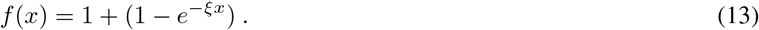

The last term of the exponents in Eq. (9) contains the probability that these contacts are contagious, which is proportional to 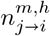, the expected number of individuals of age group *h* in the given infectious state *m* (either *A* or *I*) which have moved from region *j* to region *i*:

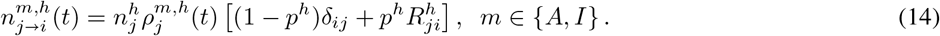

The discrete time nature of this model allows for an easy computation of the time evolution of all the relevant variables, providing information at the regional level. See Sec. IV B for the details of its application to the COVID-19 outbreak in Spain. Additionally, the model is amenable for analytical inspection, which has allowed us to find the epidemic threshold, see Appendix A.

## III. PREDICTION OF INCIDENCE UNDER MOBILITY RESTRICTIONS

Here we assess the performance of different containment measures to reduce the impact of COVID-19 using the mathematical model. To incorporate containment policies in our formalism, we assume that a given fraction of the population *κ*_0_ is isolated at home. In this sense, let us remark that parameter *κ*_0_ allows us to change the level of resolution while studying the propagation of COVID-19. Namely, with *κ*_0_ = 0 we recover the well-mixing assumption within the same municipality described in previous sections —since active population movements promote the interaction between members from different households— whereas *κ*_0_ = 1 isolates the households from each other, thus constraining the transmission dynamics at the level of household rather than municipality. From the former assumptions, we compute the average number of contacts of agents belonging to each group *g* as

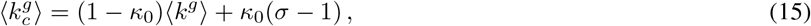

where the second term in the *r*.*h*.*s*. encodes those contacts occurring within the household, whose size (number of individuals) is assumed to be *σ* in average.

In this scenario, a relevant indicator to quantify the efficiency of the policy is the probability of one individual living in a household, inside a given municipality *i*, without any infected individual. Assuming that containment is implemented at time *t*_*c*_, this quantity, denoted in the following as *CH*_*i*_(*t*_*c*_), is given by

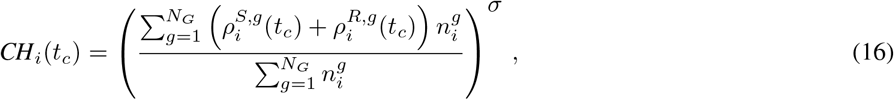

and Eq. (15) becomes time-dependent:

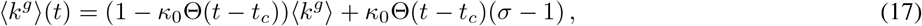

where Θ(*x*) is the Heaviside function, which is 1 if *x* ⩾ 0 and 0 otherwise. Accordingly, the mobility parameters *p*^*g*^ change as

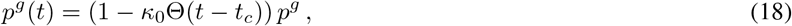

which make 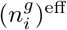 and *z*^*g*^ also dependent on time, see Eqs. (10)–(12).

This containment strategy is introduced in the dynamical Eqs. (1)–(6) by modifying Eqs. (1) and (2) for the time after *t*_*c*_:

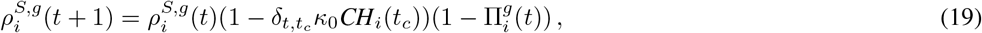

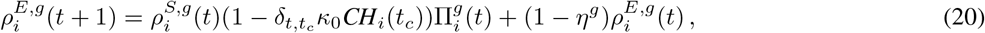

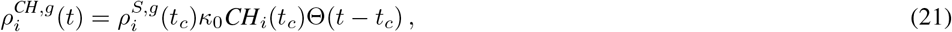

where we have added a new compartment *CH* to hold the individuals under household isolation after applying containment *κ*_0_, and *δ*_*a,b*_ is the Kronecker function, which is 1 if *a* = *b* and 0 otherwise. Containment also affects the average number of contacts, thus we must also update Eq. (9):

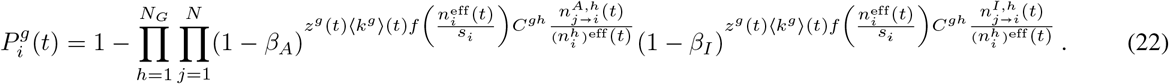

## IV. RESULTS

### A. Parameters for the modelization of the spreading of COVID-19

In this subsection, we detail our parameters choice to study the current epidemic outbreak in Spain. Regarding epidemiological parameters, the incubation period has been reported to be *η*^−1^ + *α*^−1^ = 5.2 days [2] in average which, in our formalism, must be distributed into the exposed and asymptomatic compartments. In principle, if one does not expect asymptomatic transmissions, most of this time should be spent inside the exposed compartment, thus being the asymptomatic infectious compartment totally irrelevant for disease spreading. However, along the line of several recent works [34–36] we have found that the unfolding of COVID-19 cannot be explained without accounting for infections from individuals not developing any symptoms previously. In particular, our best fit to reproduce the evolution of the real cases reported so far in Spain yields *α*^−1^ = 2.86 days as asymptomatic infectious period. In turn, the infection period is established as *µ*^−1^ = 3.2 days [1, 12], except for the young strata, for which we have reduced it to 1 day, assigning the remaining 2.2 days as asymptomatic; this is due to the reported mild symptoms in young individuals, which may become inadvertent [30]. We fix the fatality rate *ω* = 42% of ICU patients by studying historical records of dead individuals as a function of those requiring intensive care. In turn, we estimate the period from ICU admission to death as *ψ*^−1^ = 7 days [37] and the stay in ICU for those overcoming the disease as *χ*^−1^ = 10 days [38].

Regarding the population structure in Spain, we have obtained the population distribution, population pyramid, daily population flows and average household size at the municipality level from Instituto Nacional de Estadística [39] whereas the age-specific contact matrices have been extracted from [40].

### B. Prediction of the evolution of COVID-19 in Spain

Equations (1)–(7) enable to monitor the spatio-temporal propagation of COVID-19 across Spain. To check the validity of our formalism, we aggregate the number of cases predicted for each municipality at the level of autonomous regions (*comunidades autónomas*), which is a first-level political and administrative division, and compare them with the number of cases daily reported by the Spanish Health Ministry. In this sense, we compute the number of cases predicted for each municipality *i* at each time step *t* as:

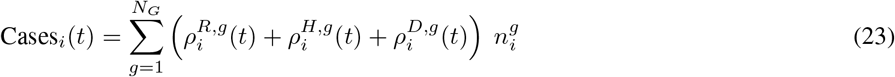

As our model is designed to predict the emergence of autochthonous cases triggered by local contagions and commuting patterns, those imported infected individuals corresponding to the first reported cases in Spain are initially plugged into our model as asymptomatic infectious agents. In addition, small infectious seeds should be also placed in those areas where anomalous outbreaks have occurred due to singular events such as one funeral in Vitoria leading to more than 60 contagions. Overall, the total number of infectious seeds is 47 individuals which represents 0.2 % of the number of cases reported by March 20, 2020.

Figure 2 shows that our model is able to accurately predict not only the overall evolution of the total number of cases at the national scale but also their spatial distribution across the different autonomous regions. Moreover, the most typical trend observed so far is an exponential growth of the number of cases, thus clearly suggesting that the disease is spreading freely in most of the territories. Note, however, that there are some exceptions such as La Rioja or País Vasco in which some strong policies targeting the most affected areas were previously promoted to slow down COVID-19 propagation.

**FIG 2.**
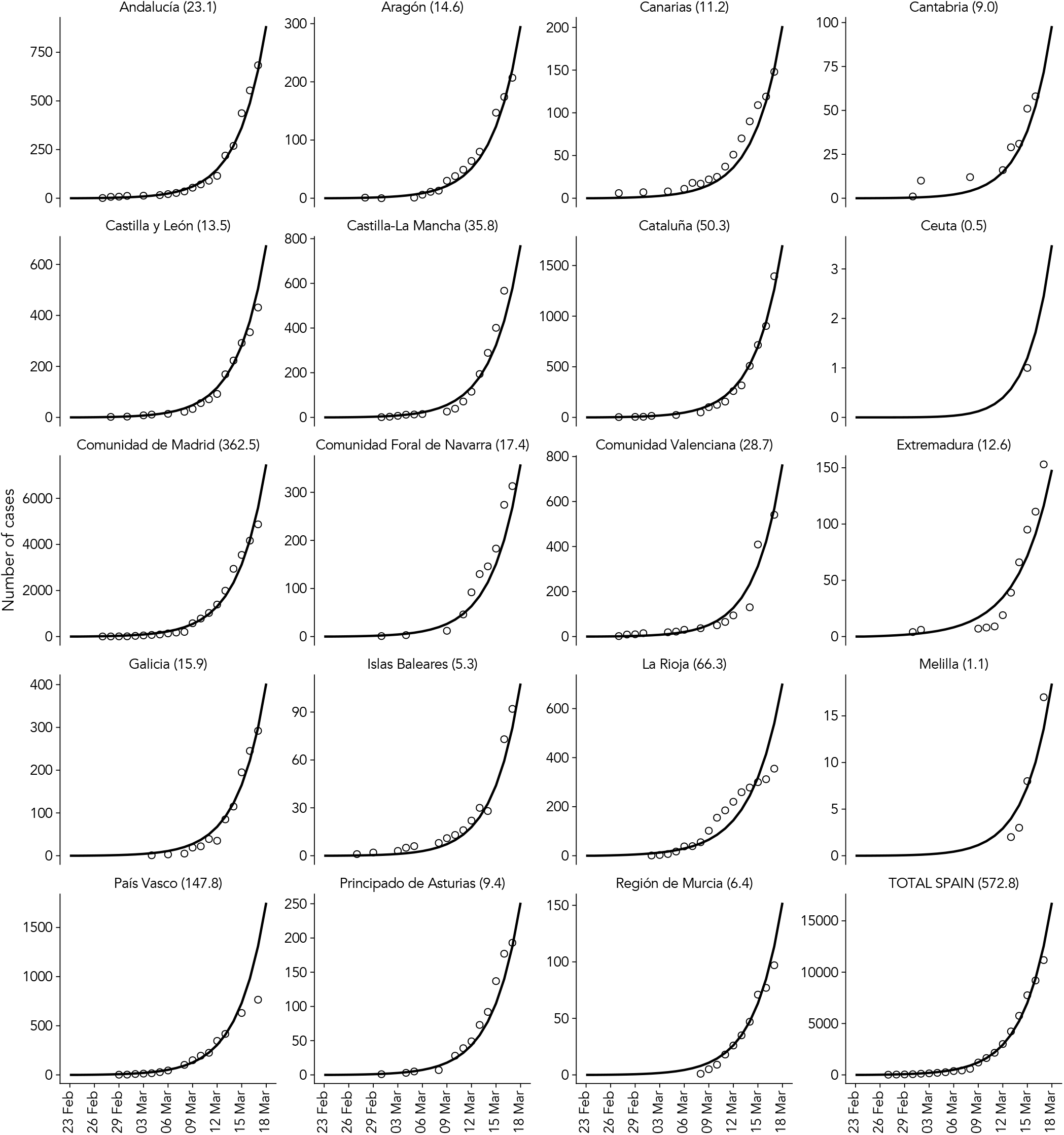
Comparison of the results of the model Eqs. (1)–(7) for each autonomous region in Spain. The solid line is the result of the epidemic model, aggregated by ages, for the number of individuals inside compartments (H+R+D) that corresponds to the expected number of cases (see Figure 1), and dots correspond to real cases reported. The number appearing next to the region name corresponds to the Mean Absolute Error (MAE) between the model prediction and the total number of cases.

To assess the impact of containment policies, we now theoretically study the effects of tuning the isolation rate *κ*_0_ controlling the fraction of population staying at home. Figure 3 shows the temporal evolution of the individuals requiring intensive care units while applying the isolation policy by March 20, 2020. Interestingly, it becomes clear that there are two different regimes. For small *κ*_0_ values, the observed behavior corresponds to the flattening of the epidemic curve while promoting social distancing. This way, increasing *κ*_0_ leads to longer epidemic periods with much less impact within society in terms of hospitalized agents. In contrast, for large enough *κ*_0_ values, the effective isolation of households allows for reducing at the same time the epidemic size and the duration of the epidemic wave. This is mainly caused by the depletion [41] of susceptible individuals which prevents the infectious individuals from sustaining the outbreak by infecting healthy peers.

**FIG 3.**
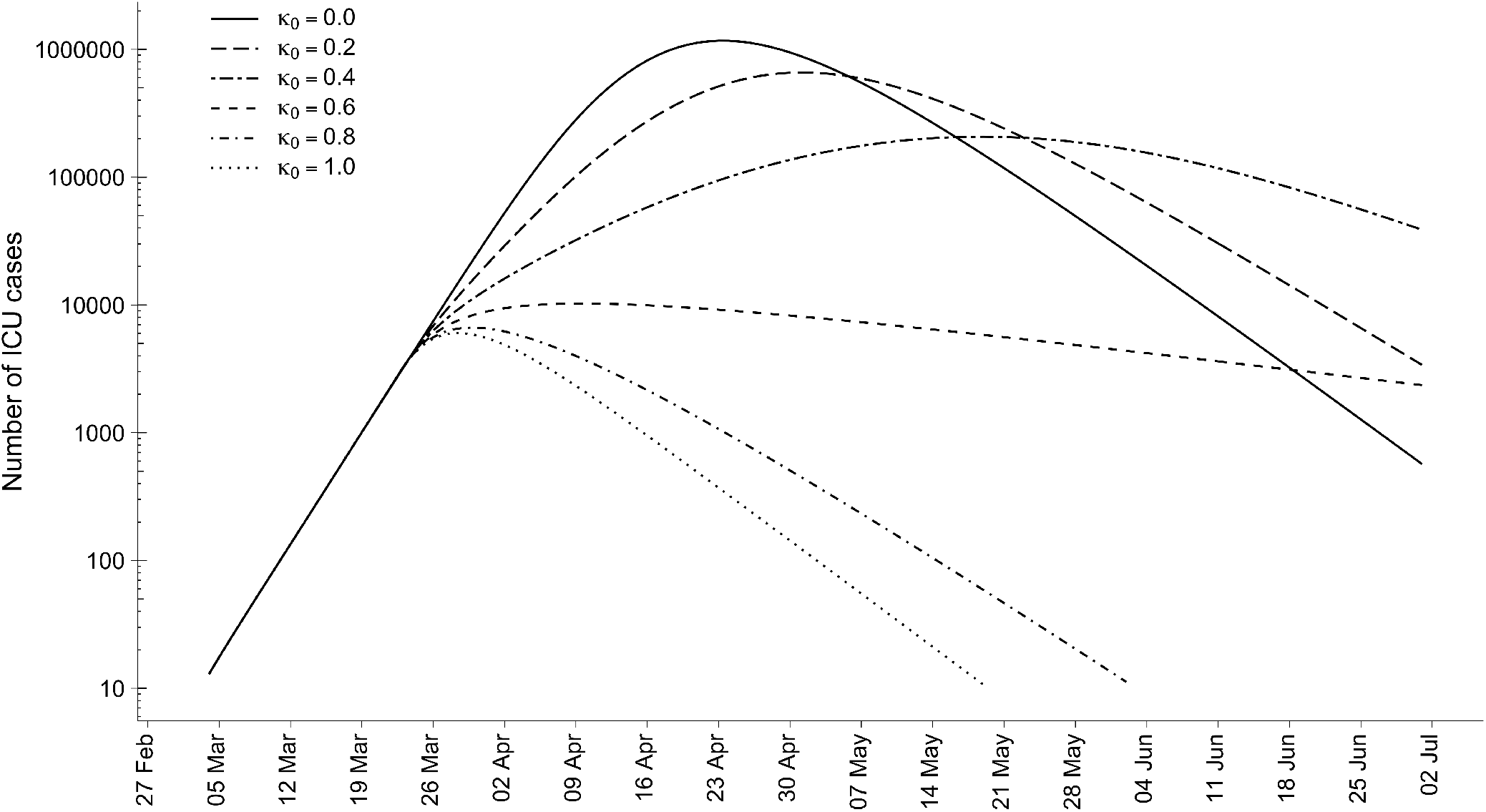
Temporal evolution of the total number of ICU cases predicted for Spain as a function of the fraction of isolated population *κ*_0_ from March 20, 2020.

Finally, we address the important health problem arising from the saturation of ICU beds. For this purpose, we study the evolution of individuals requiring intensive care units by fixing *κ*_0_ = 0.80 from March 20, 2020. To quantify the overload of ICU capacity, in Figure 4 we compare the predictions yielded by our equations with the total number of beds within each autonomous region which we estimate as the 3% of the total number of hospital beds. There we find that the saturation of hospitals across Spain is not uniform but strongly depends on both the current extension of the outbreak and the available resources in each autonomous region.

**FIG 4.**
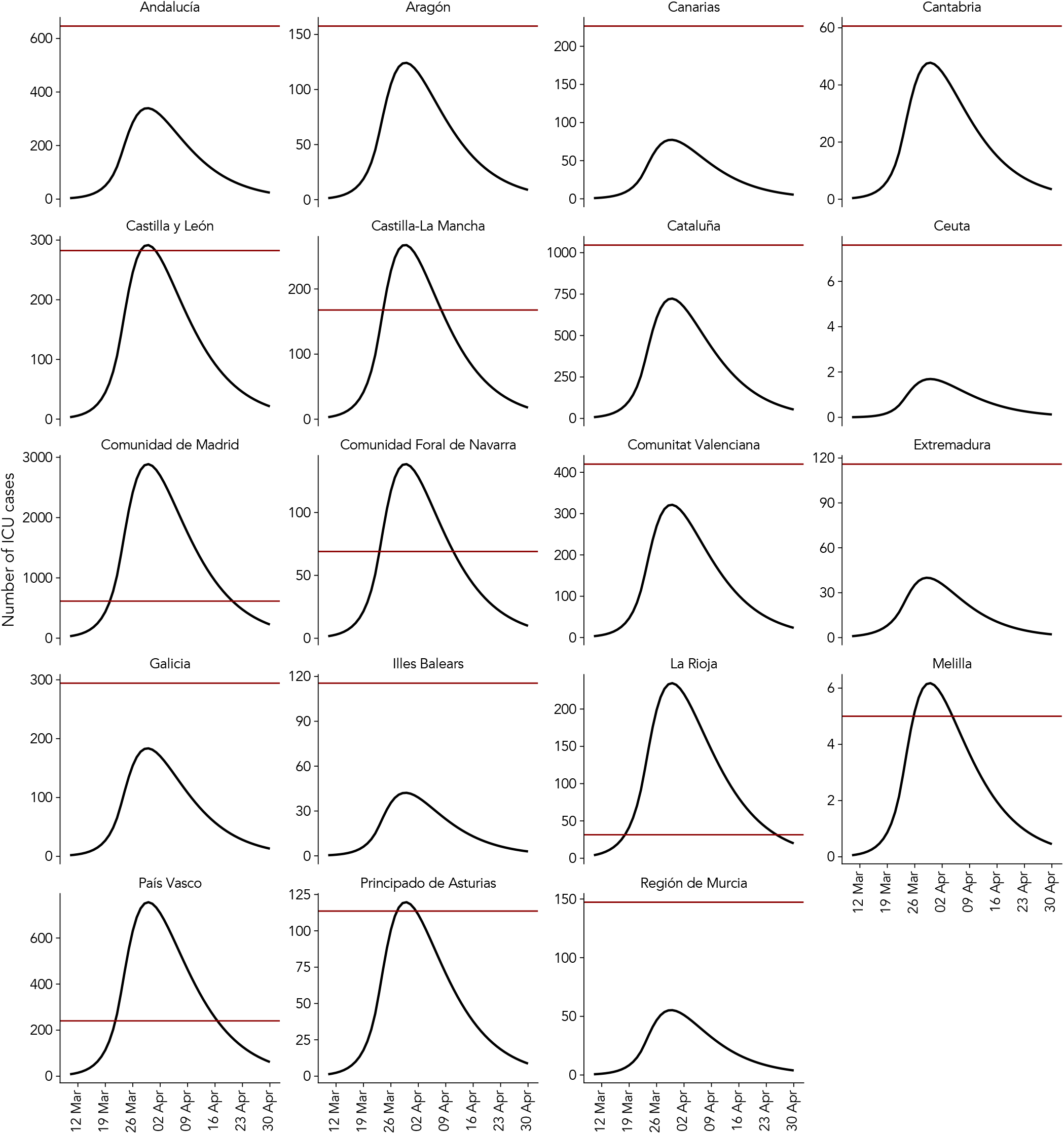
ICU saturation curves for each region in Spain. The black lines shows the temporal evolution of individuals requiring ICU. The isolation of population is performed from March 20, 2020 with *κ*_0_ = 0.80. The red line shows the estimated number of ICU beds for each autonomous region.

## V. DISCUSSION

We have presented a mathematical model based on a Microscopic Markov Chain Approach (MMCA) for the spatio-temporal spreading of COVID–19. The model captures human behavior features such as: the urban demography, age strata, age-structured contact patterns, and daily recurrent mobility flows. Importantly, the epidemiological and human characteristics present in this model provides with the possibility of a rapid and reliable evaluation of different containment policies.

We have applied the results to the validation and projection of the propagation of COVID–19 in Spain. Our results reveal that, at the current stage of the epidemics, the application of stricter containment measures of social distance are urgent to avoid the collapse of the health system. Moreover, we are close to an scenario in which the complete lockdown appears as the only possible measure to avoid the former situation. Other scenarios can be prescribed and analyzed after lockdown, as for example pulsating open-closing strategies or targeted herd immunity.

## Data Availability

All data will be available upon request.

## Appendix A Calculation of epidemic threshold

The model is amenable for analytical calculations. We calculate the epidemic threshold using the next generation matrix approach [42]. Accordingly, we need to analyze the stability of the disease free equilibrium. We do so by making a first order expansion of the above equations for small values *ϵ* of the non-susceptible states 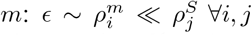 and 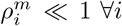, where *m* ∈ {*E, A, I, H, D, R*} The expansion allows us to transform our discrete time Markov Chain into a continuous time differential equation. We start by expanding the infection probabilities 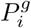:

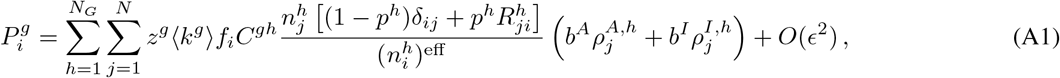

where we have defined

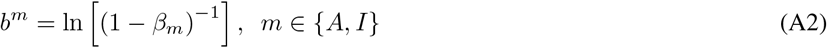

and

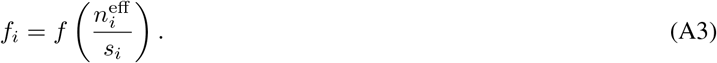

We then insert the above expression into 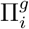, leading to:

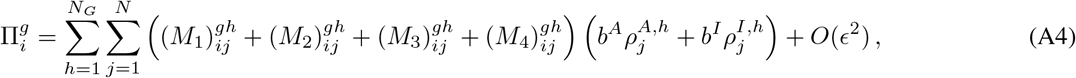

where the above tensors *M*_*ℓ*_ for *ℓ* ∈ {1, …, 4} are defined as:

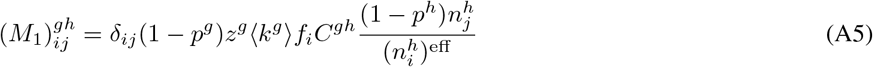

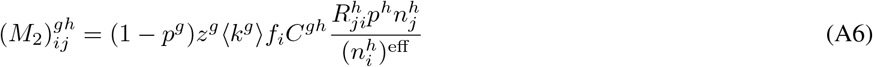

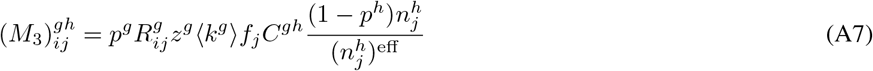

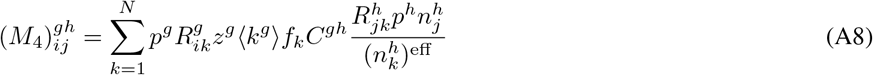

These tensors encode the four different ways in which the epidemic interactions may take place: individuals belonging to the same patch *i* = *j* and not moving (*M*_1_); interaction in the patch of *i* with individuals coming from patch *j* (*M*_2_); interaction in the patch of *j* with individuals coming from patch *i* (*M*_3_); and individuals from *i* and *j* interacting at any other patch *k* (*M*_4_).

In the next generation matrix framework, we only need to consider the epidemic compartments. Making use of the above definitions, the corresponding differential equations take the form:

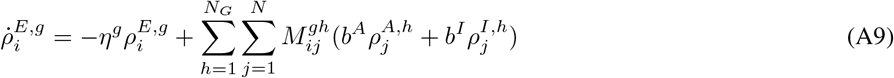

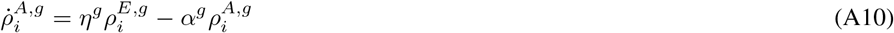

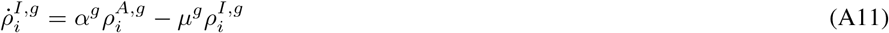

Where the tensor *M* is given by 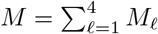 Defining the vector (*ρ*^*g*^)^*T*^ = (*ρ*^*E,g*^,*ρ*^*A,g*^ *ρ*^*I,g*^), the above system of differential equations can be rewritten as:

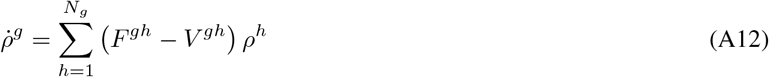

Where we defined *V* ^*gh*^ = *V* ^*g*^*δ*^*gh*^ ⊗ 𝟙_*N*×*N*_ with:

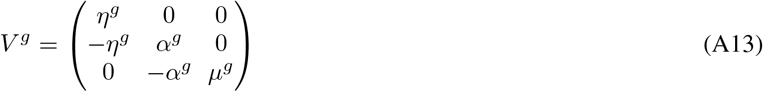

And:

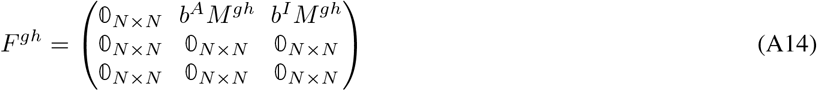

With the above differential equation, the reproduction number is given by:

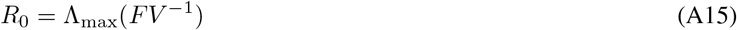

We can calculate the inverse of the tensor *V* as (*V*^−1^)^*gh*^ = (*V* ^*g*^)^−1^*δ*^*gh*^ ⊗ 𝟙_*N*×*N*_. The inverse of the matrix *V* ^*g*^ is given by:

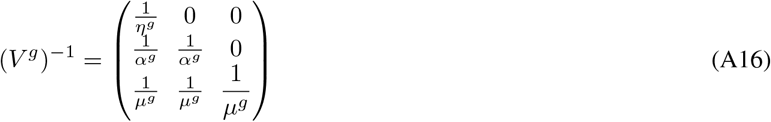

Accordingly, we have:

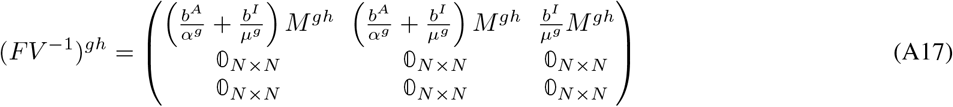

As we look for the eigenvectors of the tensor *FV* ^−1^, we note that their components associated to the compartments *A* and *I* — rows 2 and 3— must be zero, since the associated rows in the above matrix are zero. To be more precise, we have 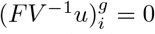 for *i* = 2*N* + 1, …3*N*, which are the elements associated to the compartments *A* and *I*. Accordingly, we can restrict the above matrix only to the vector space associated to the compartment *E* and the eigenvalues will be equivalent, which gives us:

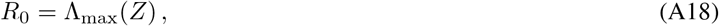

Where 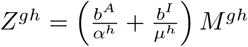. Finally, the epidemic threshold is found by solving the implicit equation Λ_max_ (*Z*) = 1.

## ACKNOWLEDGEMENTS

We thank Gourab Ghoshal and Silvio Ferreira for useful discussions. A.A., B.S. and S.G. acknowledge financial support from Spanish MINECO (grant PGC2018-094754-B-C21), Generalitat de Catalunya (grant No. 2017SGR-896), and Universitat Rovira i Virgili (grant No. 2018PFR-URV-B2-41). A.A. also acknowledge support from Generalitat de Catalunya ICREA Academia, and the James S. McDonnell Foundation grant #220020325. J.G.G. and D.S.P. acknowledges financial support from MINECO (projects FIS2015-71582-C2 and FIS2017-87519-P) and from the Departamento de Industria e Innovación del Gobierno de Aragón y Fondo Social Europeo (FENOL group E-19). C.G. acknowledges financial support from Juan de la Cierva-Formación (Ministerio de Ciencia, Innovación y Universidades). B.S. acknowledges financial support from the European Union’s Horizon 2020 research and innovation program under the Marie Sklodowska-Curie grant agreement No. 713679 and from the Universitat Rovira i Virgili (URV). W.C. acknowledges financial support from the CoordenaÇão de AperfeiÇoamento de Pessoal de Nível Superior, Brasil (CAPES), Finance Code 001.

## References

[1] Jonathan M Read, Jessica RE Bridgen, Derek AT Cummings, Antonia Ho, and Chris P Jewell. Novel coronavirus 2019-nCoV: early estimation of epidemiological parameters and epidemic predictions. medRxiv, page 2020.01.23.20018549, jan 2020.

[2] Qun Li, Xuhua Guan, Peng Wu, Xiaoye Wang, Lei Zhou, Yeqing Tong, Ruiqi Ren, Kathy S.M. Leung, Eric H.Y. Lau, Jessica Y. Wong, Xuesen Xing, Nijuan Xiang, Yang Wu, Chao Li, Qi Chen, Dan Li, Tian Liu, Jing Zhao, Man Liu, Wenxiao Tu, Chuding Chen, Lianmei Jin, Rui Yang, Qi Wang, Suhua Zhou, Rui Wang, Hui Liu, Yinbo Luo, Yuan Liu, Ge Shao, Huan Li, Zhongfa Tao, Yang Yang, Zhiqiang Deng, Boxi Liu, Zhitao Ma, Yanping Zhang, Guoqing Shi, Tommy T.Y. Lam, Joseph T. Wu, George F. Gao, Benjamin J. Cowling, Bo Yang, Gabriel M. Leung, and Zijian Feng. Early Transmission Dynamics in Wuhan, China, of Novel Coronavirus–Infected Pneumonia. New England Journal of Medicine, jan 2020.

[3] Yang Yang, Qingbin Lu, Mingjin Liu, Yixing Wang, Anran Zhang, Neda Jalali, Natalie Dean, Ira Longini, M. Elizabeth Halloran, Bo Xu, Xiaoai Zhang, Liping Wang, Wei Liu, and Liqun Fang. Epidemiological and clinical features of the 2019 novel coronavirus outbreak in China. medRxiv, page 2020.02.10.20021675, feb 2020.

[4] Wei-jie Guan, Zheng-yi Ni, Yu Hu, Wen-hua Liang, Chun-quan Ou, Jian-xing He, Lei Liu, Hong Shan, Chun-liang Lei, David SC Hui, Bin Du, Lan-juan Li, Guang Zeng, Kowk-Yung Yuen, Ru-chong Chen, Chun-li Tang, Tao Wang, Ping-yan Chen, Jie Xiang, Shi-yue Li, Jin-lin Wang, Zi-jing Liang, Yi-xiang Peng, Li Wei, Yong Liu, Ya-hua Hu, Peng Peng, Jian-ming Wang, Ji-yang Liu, Zhong Chen, Gang Li, Zhi-jian Zheng, Shao-qin Qiu, Jie Luo, Chang-jiang Ye, Shao-yong Zhu, and Nan-shan Zhong. Clinical characteristics of 2019 novel coronavirus infection in China. medRxiv, page 2020.02.06.20020974, feb 2020.

[5] Julien Riou and Christian L. Althaus. Pattern of early human-to-human transmission of Wuhan 2019 novel coronavirus (2019-nCoV), December 2019 to January 2020. Euro surveillance : bulletin Europeen sur les maladies transmissibles = European communicable disease bulletin, 25(4), 2020.

[6] Zhidong Cao, Qingpeng Zhang, Xin Lu, Dirk Pfeiffer, Zhongwei Jia, Hongbing Song, and Daniel Dajun Zeng. Estimating the effective reproduction number of the 2019-nCoV in China. medRxiv, page 2020.01.27.20018952, 2020.

[7] Tang, Wang, Li, Bragazzi, Tang, Xiao, and Wu. Estimation of the Transmission Risk of the 2019-nCoV and Its Implication for Public Health Interventions. Journal of Clinical Medicine, 9(2):462, 2020.

[8] Steven Sanche, Yen Ting Lin, Chonggang Xu, Ethan Romero-Severson, Nicolas W. Hengartner, and Ruian Ke. The Novel Coronavirus, 2019-nCoV, is Highly Contagious and More Infectious Than Initially Estimated. feb 2020.

[9] Jantien A. Backer, Don Klinkenberg, and Jacco Wallinga. Incubation period of 2019 novel coronavirus (2019-nCoV) infections among travellers from Wuhan, China, 20-28 January 2020. Euro surveillance : bulletin Europeen sur les maladies transmissibles = European communicable disease bulletin, 25(5), 2020.

[10] Lauren Tindale, Michelle Coombe, Jessica E Stockdale, Emma Garlock, Wing Yin Venus Lau, Manu Saraswat, Yen-Hsiang Brian Lee, Louxin Zhang, Dongxuan Chen, Jacco Wallinga, and Caroline Colijn. Transmission interval estimates suggest pre-symptomatic spread of COVID-19. medRxiv, page 2020.03.03.20029983, 2020.

[11] Matteo Chinazzi, Jessica T. Davis, Marco Ajelli, Corrado Gioannini, Maria Litvinova, Stefano Merler, Ana Pastore y Piontti, Luca Rossi, Kaiyuan Sun, Cécile Viboud, Xinyue Xiong, Hongjie Yu, M. Elizabeth Halloran, Ira M. Longini, and Alessandro Vespignani. The effect of travel restrictions on the spread of the 2019 novel coronavirus (2019-nCoV) outbreak. medRxiv, page 2020.02.09.20021261, feb 2020.

[12] Leon Danon, Ellen Brooks-Pollock, Mick Bailey, and Matt J Keeling. A spatial model of CoVID-19 transmission in England and Wales: early spread and peak timing. medRxiv, page 2020.02.12.20022566, feb 2020.

[13] Joseph T. Wu, Kathy Leung, and Gabriel M. Leung. Nowcasting and forecasting the potential domestic and international spread of the 2019-nCoV outbreak originating in Wuhan, China: a modelling study. The Lancet, 395(10225):689–697, feb 2020.

[14] Neil M. Ferguson, Daniel Laydon, Gemma Nedjati-Gilani, Natsuko Imai, Kylie Ainslie, Marc Baguelin, Sangeeta Bhatia, Adhiratha Boonyasiri, Zulma Cucunubá, Gina Cuomo-Dannenburg, Amy Dighe, Ilaria Dorigatti, Han Fu, Katy Gaythorpe, Will Green, Arran Hamlet, Wes Hinsley, Lucy C Okell, Sabine Van Elsland, Hayley Thompson, Robert Verity, Erik Volz, Haowei Wang, Yuanrong Wang, Patrick Gt Walker, Caroline Walters, Peter Winskill, Charles Whittaker, Christl A Donnelly, Steven Riley, and Azra C Ghani. Impact of non-pharmaceutical interventions (NPIs) to reduce COVID-19 mortality and healthcare demand.

[15] Marius Gilbert, Giulia Pullano, Francesco Pinotti, Eugenio Valdano, Chiara Poletto, Pierre Yves Boëlle, Eric D’Ortenzio, Yazdan Yaz-danpanah, Serge Paul Eholie, Mathias Altmann, Bernardo Gutierrez, Moritz U.G. Kraemer, and Vittoria Colizza. Preparedness and vulnerability of African countries against importations of COVID-19: a modelling study. The Lancet, 2020.

[16] Giulia Pullano, Francesco Pinotti, Eugenio Valdano, Pierre Yves Boëlle, Chiara Poletto, and Vittoria Colizza. Novel coronavirus (2019- nCoV) early-stage importation risk to Europe, January 2020. Euro surveillance : bulletin Europeen sur les maladies transmissibles = European communicable disease bulletin, 25(4), 2020.

[17] Juanjuan Zhang, Maria Litvinova, Wei Wang, Yan Wang, Xiaowei Deng, Xinghui Chen, Mei Li, Wen Zheng, Lan Yi, Xinhua Chen, Qianhui Wu, Yuxia Liang, Xiling Wang, Juan Yang, Kaiyuan Sun, Ira M. Longini, M. Elizabeth Halloran, Peng Wu, Benjamin J. Cowling, Stefano Merler, Cecile Viboud, Alessandro Vespignani, Marco Ajelli, and Hongjie Yu. Evolving epidemiology of novel coronavirus diseases 2019 and possible interruption of local transmission outside Hubei Province in China: a descriptive and modeling study. medRxiv, page 2020.02.21.20026328, 2020.

[18] Jesús Gómez-Gardeñes, David Soriano-Paños, and Alex Arenas. Critical regimes driven by recurrent mobility patterns of reaction- diffusion processes in networks. Nature Physics, 14(4):391–395, apr 2018.

[19] D. Soriano-Paños, L. Lotero, A. Arenas, and J. Gómez-Gardeñes. Spreading Processes in Multiplex Metapopulations Containing Differ- ent Mobility Networks. Physical Review X, 8(3):031039, aug 2018.

[20] David Soriano-Paños, Gourab Ghoshal, Alex Arenas, and Jesús Gómez-Gardeñes. Impact of temporal scales and recurrent mobility patterns on the unfolding of epidemics. Journal of Statistical Mechanics: Theory and Experiment, 2020(2):024006, feb 2020.

[21] David Soriano-Paños, Juddy Heliana Arias-Castro, Adriana Reyna-Lara, Hector J. Martínez, Sandro Meloni, and Jesús Gómez-Gardeñes. Vector-borne epidemics driven by human mobility. Phys. Rev. Research, b2:013312, Mar 2020.

[22] Sergio Gómez, Alex Arenas, Javier Borge-Holthoefer, Sandro Meloni, and Yamir Moreno. Discrete-time Markov chain approach to contact-based disease spreading in complex networks. EPL (Europhysics Letters), 89(3):38009, feb 2010.

[23] Sergio Gómez, Jesús Gómez-Gardenes, Yamir Moreno, and Alex Arenas. Nonperturbative heterogeneous mean-field approach to epi- demic spreading in complex networks. Physical Review E, 84(3):036105, 2011.

[24] Clara Granell, Sergio Gómez, and Alex Arenas. Dynamical interplay between awareness and epidemic spreading in multiplex networks. Physical Review Letters, 111(12), 2013.

[25] Clara Granell, Sergio Gómez, and Alex Arenas. Competing spreading processes on multiplex networks: Awareness and epidemics. Physical Review E - Statistical, Nonlinear, and Soft Matter Physics, 90(1), 2014.

[26] Joan T Matamalas, Alex Arenas, and Sergio Gómez. Effective approach to epidemic containment using link equations in complex networks. Science Advances, 4(12):eaau4212, 2018.

[27] Joan T Matamalas, Sergio Gómez, and Alex Arenas. Abrupt phase transition of epidemic spreading in simplicial complexes. Physical Review Research, 2(1):012049, 2020.

[28] Stephanie Bialek, Ellen Boundy, Virginia Bowen, Nancy Chow, Amanda Cohn, Nicole Dowling, Sascha Ellington, Ryan Gierke, Aron Hall, Jessica MacNeil, Priti Patel, Georgina Peacock, Tamara Pilishvili, Hilda Razzaghi, Nia Reed, Matthew Ritchie, and Erin Sauber-Schatz. Severe Outcomes Among Patients with Coronavirus Disease 2019 (COVID-19) — United States, February 12–March 16, 2020. MMWR. Morbidity and Mortality Weekly Report, 69(12), mar 2020.

[29] Articles Clinical course and risk factors for mortality of adult inpatients with COVID-19 in Wuhan, China : a retrospective cohort study. The Lancet, 6736(20):1–9, 2020.

[30] Qifang Bi, Yongsheng Wu, Shujiang Mei, Chenfei Ye, Xuan Zou, Zhen Zhang, Xiaojian Liu, Lan Wei, Shaun A Truelove, Tong Zhang, Wei Gao, Cong Cheng, Xiujuan Tang, Xiaoliang Wu, Yu Wu, Binbin Sun, Suli Huang, Yu Sun, Juncen Zhang, Ting Ma, Justin Lessler, and Teijian Feng. Epidemiology and transmission of covid-19 in shenzhen china: Analysis of 391 cases and 1,286 of their close contacts. medRxiv, 2020.

[31] Clara Granell and Peter J. Mucha. Epidemic spreading in localized environments with recurrent mobility patterns. Physical Review E, 97(5), 2018.

[32] Albert Solé-Ribalta, Sergio Gómez, and Alex Arenas. A model to identify urban traffic congestion hotspots in complex networks. Royal Society Open Science, 3(10), 2016.

[33] Hao Hu, Karima Nigmatulina, and Philip Eckhoff. The scaling of contact rates with population density for the infectious disease models. Mathematical Biosciences, 244(2):125–134, 2013.

[34] Ruiyun Li, Sen Pei, Bin Chen, Yimeng Song, Tao Zhang, Wan Yang, and Jeffrey Shaman. Substantial undocumented infection facilitates the rapid dissemination of novel coronavirus (sars-cov2). Science, 2020.

[35] Hiroshi Nishiura, Natalie M. Linton, and Andrei R. Akhmetzhanov. Serial interval of novel coronavirus (covid-19) infections. International Journal of Infectious Diseases, 2020.

[36] Tapiwa Ganyani, Cecile Kremer, Dongxuan Chen, Andrea Torneri, Christel Faes, Jacco Wallinga, and Niel Hens. Estimating the generation interval for covid-19 based on symptom onset data. medRxiv, 2020.

[37] Nick Wilson, Amanda Kvalsvig, Lucy Telfar Barnard, and Michael G. Baker. Case-fatality risk estimates for covid-19 calculated by using a lag time for fatality. 26(6), 2020.

[38] Gemma Nedjati-Gilani Natsuko Imai Kylie Ainslie Marc Baguelin Sangeeta Bhatia Adhiratha Boonyasiri Zulma Cucunubá Gina Cuomo-Dannenburg Amy Dighe Ilaria Dorigatti Han Fu Katy Gaythorpe Will Green Arran Hamlet Wes Hinsley Lucy C Okell Sabine van Elsland Hayley Thompson Robert Verity Erik Volz Haowei Wang Yuanrong Wang Patrick GT Walker Caroline Walters Peter Winskill Charles Whittaker Christl A Donnelly Steven Riley Azra C Ghani Neil M. Ferguson, Daniel Laydon. Impact of non-pharmaceutical interventions (npis) to reduce covid- 19 mortality and healthcare demand. Imperial College COVID-19 Response Team, Mar 2020.

[39] Instituto Nacional de Estadistica. spain. https://www.ine.es. Accessed: 2020-02-26.

[40] Kiesha Prem, Alex R. Cook, and Mark Jit. Projecting social contact matrices in 152 countries using contact surveys and demographic data. PLOS Computational Biology, 13(9):1–21, 09 2017.

[41] Benjamin F. Maier and Dirk Brockmann. Effective containment explains sub-exponential growth in confirmed cases of recent covid-19 outbreak in mainland china, 2020.

[42] O. Diekmann, J. A.P. Heesterbeek, and M. G. Roberts. The construction of next-generation matrices for compartmental epidemic models. Journal of the Royal Society Interface, 7(47):873–885, 2010.

